# ASSOCIATION OF HYPERGLYCEMIA WITH HOSPITAL MORTALITY IN NONDIABETIC COVID-19 PATIENTS: A COHORT STUDY

**DOI:** 10.1101/2020.08.31.20185157

**Authors:** Manju Mamtani, Ambarish M. Athavale, Mohan Abraham, Jane Vernik, Amatur R Amarah, Juan P. Ruiz, Amit J. Joshi, Mathew Itteera, Sara D Zhukovski, Ravi Prakash Madaiah, Blaine C White, Peter Hart, Hemant Kulkarni

**Affiliations:** M&H Research, LLC, San Antonio, Texas, USA; Division of Nephrology, Department of Medicine, Cook County Health, Chicago, Illinois, USA; Rush Medical College, Chicago, Illinois, USA; Cerner Corporation, Kansas City, Missouri, USA; Department of Emergency Medicine, Wayne State University School of Medicine, Detroit, Michigan, USA

## Abstract

**Objective:** Diabetes is a known risk factor for mortality in Coronavirus disease 2019 (COVID-19) patients. Our objective was to identify prevalence of hyperglycemia in COVID-19 patients with and without prior diabetes and quantify its association with COVID-19 disease course.

**Research Design and Methods:** This observational cohort study included all consecutive COVID-19 patients admitted to John H Stroger Jr. Hospital, Chicago, IL from March 15, 2020 to May 15, 2020. The primary outcome was hospital mortality and the studied predictor was hyperglycemia (any blood glucose ≥7.78 mmol/L during hospitalization).

**Results:** Of 403 COVID-19 patients studied, 51 (12.7%) died. Hyperglycemia occurred in 228 (57%) patients; 83 of these hyperglycemic patients (36%) had no prior history of diabetes. Compared to the reference group no-diabetes/no-hyperglycemia patients the no-diabetes/hyperglycemia patients showed higher mortality [1.8% versus 20.5%, adjusted odds ratio 21.94 (95% confidence interval 4.04-119.0), p < 0.001]; improved prediction of death (p=0.0162) and faster progression to death (p=0.0051). Hyperglycemia within the first 24 and 48 hours was also significantly associated with mortality (odds ratio 2.15 and 3.31, respectively). Further, compared to the same reference group, the no-diabetes/hyperglycemia patients had higher likelihood of ICU admission (p<0.001), acute respiratory distress syndrome (p<0.001), mechanical ventilation (p<0.001), and a longer hospital stay in survivors (p<0.001).

**Conclusions:** Hyperglycemia without prior diabetes was common (21% of hospitalized COVID-19 patients) and was associated with an increased risk of and faster progression to death. Development of hyperglycemia in COVID-19 patients who do not have diabetes is an early indicator of progressive disease.

## INTRODUCTION

The coronavirus disease 2019 (COVID-19) pandemic has resulted in over 48 million cases and 1.2 million deaths globally[1]. Diabetes is associated with a higher mortality, need for intensive care, acute respiratory distress syndrome in COVID-19 disease.[2] Diabetes (HbA1_C_ ≥6.5%) and/or uncontrolled hyperglycemia (≥2 glucose measurements >10.0 mmol/L) are associated with poor outcomes in COVID-19 patients.[3] Stress hyperglycemia (defined as blood glucose values exceeding 7.78 mmol/L) in the absence of diabetes is seen in severe acute illness[4, 5].[6] Previous studies have shown that in critically ill patients, stress hyperglycemia is associated with poor clinical outcomes during hospitalization.[7] Stress hyperglycemia can prolong the length of hospital stay [8, 9] – a parameter that is closely linked with poor outcomes in COVID-19 patients[10]. An excess of circulating proinflammatory cytokines (common in COVID-19 patients) is associated with occurrence and consequences of hyperglycemia.[11-13] Sardu et al [14] showed that hyperglycemia during hospitalization correlated with interleukin-6 and D-dimer concentrations in COVID-19 patients. However, whether hyperglycemia in absence of Diabetes has a potential role in COVID-19 disease course is not clear.

In this investigation, we focused on the potential of hyperglycemia detected early during hospitalization of COVID-19 as an indicator of mortality. We hypothesized that hyperglycemia even in the absence of diabetes may be associated with adverse outcomes in COVID-19 patients. Here, we report the independent association of hyperglycemia with clinical course in COVID-19 patients using a single-center data of hospitalized COVID-19 patients from the United States of America.

## METHODS

### Study participants

This study was conducted at John H. Stroger, Jr Hospital of Cook County, Chicago, IL. All COVID-19 patients admitted between March 15, 2020 and May 3, 2020 and followed till the censoring date of May 15, 2020 were included. On the censoring day, 17 (4.2%) patients were still in hospital. COVID-19 was confirmed using the polymerase chain reaction for the RdRp and N genes. Clinical data of these patients was collected by chart reviews. The study was approved by the Institutional Review Board of the Cook County Health, Chicago, IL with waiver of informed consent.

### Outcomes and predictors

The primary outcomes were hospital mortality and time to progress to mortality. Secondary outcomes were – admission to intensive care unit (ICU), mechanical ventilation, development of acute respiratory distress syndrome (ARDS) and length of hospital stay. Outcome ascertainment was censored on May 15, 2020. Patients still in hospital who did develop an outcome under consideration were censored for computation of length of stay and time-to-event analyses.

Main predictor of interest was hyperglycemia defined as at least one BG value ≥7.78 mmol/L – a cutoff recommended as a treatment target in critically ill patients [15] and a definition of hyperglycemia in non-critically ill hospitalized patients [16]. To examine the use of hyperglycemia as an early predictor of adverse outcomes, we also considered occurrence of hyperglycemia within the first 24 hours (HG_24_) and 48 hours (HG_48_) of admission. BG values were retrospectively derived from the database and represented a mixture of fasting and non-fasting measurements and venous and capillary sources. Detailed information collected on socio-demographics, presenting symptoms, comorbidities, laboratory investigations, history of medications and substance use from the electronic health records. Severity of illness at admission was quantified using the qSOFA score that combines information from respiratory rate, systolic blood pressure and mental status into a single metric.[17]

### Statistical analyses

Descriptive statistics included mean and standard deviation (for continuous variables) and proportion (for categorical variables). The association of diabetes and hyperglycemia with outcomes was tested for significance using the Pearson’s chi-square test and the Kruskal-Wallis test as appropriate. The association with mortality was quantified as odds ratios (OR) using multivariable stepwise logistic regression analyses with forward addition strategy and a retention criterion of 0.05. The forward addition strategy was used since it is robust to the number of predictors for a moderate size dataset and accounts for the potential collinearity among covariates without overfitting the data. Since some patients initially not identified as having diabetes could have been misclassified, we conducted sensitivity analyses using a three-pronged approach: a. we restricted the analyses to patients on whom HbA1C data was available; b. in this subset we reclassified patients with HbA1C ≥6.5% as diabetes; and c. We conducted bootstrapping with 1000 replicates for robust estimation of 95% confidence intervals (CI). The differential rate of progression to death was examined using the Kaplan-Meier plot and tested for significance using the logrank test. The time trends of BG values during hospitalization were examined using generalized estimating equations (GEE). The GEE models used Gaussian family function, identity link function and equal correlation structure. BG time trends were smoothed using cubic splines with a knot every day for the first 14 days of admission. Improvement in the prediction of the primary outcome using hyperglycemia was assessed by estimating the area under a receiver-operating characteristic curve (AUROC). Statistical significance for difference between two AUROCs was tested using the DeLong and DeLong test. All statistical analyses were conducted using Stata 12.0 software package (Stata Corp., College Station, TX). The statistical significance was assessed at a type I error rate of 0.05.

## RESULTS

### Study participants

We included a total of 403 (out of 406 eligible, 99.3%) COVID-19 patients who were admitted to the study center and had non-missing data for diabetes and other comorbidities. Clinical characteristics of the study participants are detailed in Table 1. Majority of the patients were male (67.7%), of Hispanic/Lati2no ethnicity (54.84%) and Black/African American race (38%). 97 (24%) patients needed ICU admission; 56 (∼14%) patients needed mechanical ventilation; and 51 (∼13%) patients died.

**Table 1.** Baseline characteristics of study participants (total n = 403)

| Characteristic† | Mean / N* | SD / % | N available** |
| --- | --- | --- | --- |
| ***Socio-demographic characteristics*** |  |  |  |
| Age (y) | 54.96 | 13.55 | 403 |
| Males | 273 | 67.74 | 403 |
| Hispanic/Latino ethnicity | 221 | 54.84 | 403 |
| Race |  |  |  |
| Black / African American | 153 | 37.97 | 403 |
| White | 137 | 34.00 | 403 |
| Other | 79 | 19.60 | 403 |
| American Indian | 25 | 6.20 | 403 |
| Asian | 7 | 1.74 | 403 |
| Multiple | 2 | 0.50 | 403 |
| ***Presenting symptoms*** |  |  |  |
| Cough | 101 | 25.06 | 403 |
| Fever | 75 | 18.61 | 403 |
| Shortness of breath | 61 | 15.14 | 403 |
| Myalgia / arthralgia | 47 | 11.66 | 403 |
| Fatigue | 35 | 8.68 | 403 |
| Fever with chills | 28 | 6.95 | 403 |
| Cough with sputum | 23 | 5.71 | 403 |
| Diarrhea | 21 | 5.21 | 403 |
| Sore throat | 10 | 2.48 | 403 |
| Nausea / vomiting | 10 | 2.48 | 403 |
| Nasal congestion | 8 | 1.99 | 403 |
| Headache | 7 | 1.74 | 403 |
| Altered mental status | 7 | 1.74 | 403 |
| Hemoptysis | 2 | 0.50 | 403 |
| ***Comorbidities*** |  |  |  |
| Hypertension | 196 | 48.64 | 403 |
| Obesity (BMI $\geq 30$ Kg/m<sup>2</sup>) | 185 | 45.91 | 403 |
| Diabetes | 155 | 38.46 | 403 |
| Coronary artery disease | 31 | 7.69 | 403 |
| Chronic kidney disease | 31 | 7.69 | 403 |
| Asthma | 27 | 6.70 | 403 |
| Other lung disease | 25 | 6.20 | 403 |
| Chronic liver disease | 25 | 6.20 | 403 |
| Cancer | 25 | 6.20 | 403 |
| Chronic heart failure | 21 | 5.21 | 403 |
| COPD | 17 | 4.22 | 403 |
| ESRD | 15 | 3.72 | 403 |
| HIV/AIDS | 10 | 2.48 | 403 |
| Atrial fibrillation | 9 | 2.23 | 403 |
| Smoking status |  |  |  |
| Non-smoker | 276 | 68.83 | 401 |
| Former smoker | 51 | 12.72 | 401 |
| Current smoker | 39 | 9.73 | 401 |
| Unknown | 35 | 8.73 | 401 |
| qSOFA score |  |  |  |
| qSOFA 0 | 110 | 27.30 | 403 |
| qSOFA 1 | 205 | 50.87 | 403 |
| qSOFA 2 | 86 | 21.34 | 403 |
| qSOFA 3 | 2 | 0.50 | 403 |
| **Other early features** |  |  |  |
| Highest temperature in 24 hours (°C) | 38.19 | 0.89 | 402 |
| Lowest systolic BP in 24 hours (mmHg) | 108.90 | 15.88 | 403 |
| Highest heart rate in 24 hours (bpm) | 105.08 | 16.53 | 403 |
| Highest respiratory rate in 24 hours (/min) | 26.96 | 13.12 | 403 |
| **Initial laboratory investigations** |  |  |  |
| White cell count (x10<sup>9</sup> cells/L) | 7.56 | 3.97 | 402 |
| Platelet count (x10<sup>9</sup> cells/L) | 224.15 | 112.90 | 402 |
| Hemoglobin (g/L) | 1.34 | 0.22 | 402 |
| Differential white cell count |  |  |  |
| Neutrophils (%) | 74.12 | 11.66 | 374 |
| Lymphocytes (%) | 16.58 | 9.29 | 373 |
| Eosinophils (%) | 0.43 | 0.99 | 373 |
| Basophils (%) | 0.45 | 0.30 | 375 |
| Monocytes (%) | 8.42 | 3.68 | 373 |
| Serum ferritin (µg/L) | 784.88 | 1063.95 | 263 |
| Serum sodium (mEq/L) | 135.39 | 5.00 | 403 |
| Serum potassium (mEq/L) | 4.15 | 0.59 | 371 |
| Serum bicarbonates (mEq/L) | 24.10 | 4.17 | 403 |
| First blood glucose level (mmol/L) | 8.04 | 4.24 | 403 |
| Serum creatinine (µmol/L) | 145.01 | 221.93 | 403 |
| Serum albumin (g/L) | 0.35 | 0.05 | 375 |
| Serum globulin (g/L) | 0.30 | 0.10 | 403 |
| Serum AST (U/L) | 55.24 | 78.39 | 357 |
| Serum ALT (IU/L) | 44.74 | 55.53 | 375 |
| Serum LDH (U/L) | 375.84 | 475.00 | 313 |
| Serum D-dimer (mg/L) | 2.18 | 3.30 | 221 |
| Lowest plasminogen (mg/L) | 540.36 | 190.66 | 152 |
| Serum Troponin (µg/L) | 0.26 | 1.86 | 116 |
| Serum creatine kinase (U/L) | 2682.92 | 16006.10 | 53 |
| Serum C-reactive protein (mg/L) | 12.72 | 8.88 | 279 |
| Proteinuria | 8 | 1.99 | 403 |
| Hematuria | 52 | 12.90 | 403 |
| HbA1c (%) | 7.22 | 2.29 | 279 |
| **Medication history** |  |  |  |
| Insulin | 69 | 46.00 | 150 |
| ACE inhibitors | 85 | 21.09 | 403 |
| Angiotensin receptor blockers | 28 | 6.95 | 403 |
| Mineralocorticoid receptor antagonist | 10 | 2.48 | 403 |
| Beta-blocker | 62 | 15.38 | 403 |
| Other antihypertensive | 103 | 25.56 | 403 |
| Statin | 137 | 34.00 | 403 |
| NSAID | 28 | 6.95 | 403 |
| Aspirin | 77 | 19.11 | 403 |
| Vitamin C supplementation | 1 | 0.25 | 403 |
| Vitamin D supplementation | 6 | 1.49 | 403 |
| Hydroxychloroquine | 2 | 0.50 | 403 |
| Azithromycin | 4 | 0.99 | 403 |
| Warfarin | 10 | 2.48 | 403 |
| Apixiban | 4 | 0.99 | 403 |
| Riveroxaban | 7 | 1.74 | 403 |
| Steroids | 3 | 0.74 | 403 |
| Chemotherapy | 16 | 3.97 | 403 |
| Calcineurin inhibitor | 3 | 0.74 | 403 |
| Mycophenolate mofetil | 2 | 0.50 | 403 |
| Azathioprine | 1 | 0.25 | 403 |
| **Substance use** |  |  |  |
| Marijuana | 16 | 3.97 | 403 |
| Cocaine | 19 | 4.71 | 403 |
| Heroin | 22 | 5.46 | 403 |
| Amphetamine | 2 | 0.50 | 403 |
| **Outcomes** |  |  |  |
| ICU admission | 97 | 24.06 | 403 |
| ARDS | 61 | 15.14 | 403 |
| Mechanical ventilation | 56 | 13.90 | 403 |
| Death | 51 | 12.66 | 403 |
| Length of stay (d) | 7.29 | 7.35 | 403 |

\*, Columns indicate mean and standard deviation (SD) for continuous variables and number (N) and proportion for categorical variables
\*\*, Total number of study participants on whom data was available
†, Parentheses show units

### Prevalence of hyperglycemia and diabetes

Hyperglycemia was observed in 228 (56.6%) patients (Figure 1A, yellow and red slices of the pie combined). The presence of hyperglycemia and diabetes identified four subsets of patients: those with diabetes and hyperglycemia (DM^+^/HG^+^, n = 145, 36.0%), patients with diabetes but no hyperglycemia (DM^+^/HG^-^, n = 10, 2.5%), patients with hyperglycemia who did not have diabetes (DM^-^/HG^+^, n = 83, 20.6%) and patients who had neither diabetes nor hyperglycemia (DM^-^/HG^-^, n = 165, 40.9%) (Figure 1A). HbA1c values within the past year were available for 279 patients (69.2%, Table 1). The median (interquartile range) HbA1c values for the four groups were as follows: DM^+^/HG^+^ group (available n = 141) – 8 % (3.3%); DM^-^/HG^+^ group (available n = 51) – 5.7% (0.8%); DM^+^/HG^-^ group (available n = 8) – 6.75% (0.75%); and DM^-^/HG^-^ group (available n = 79) – 5.6% (0.6%). Three (2.1%) patients belonging to the DM^-^/HG^-^ group and 10 (19.6%) patients belonging to the DM^-^/HG^+^ group had one HbA1c value ≥6.5%.

**Figure 1.**
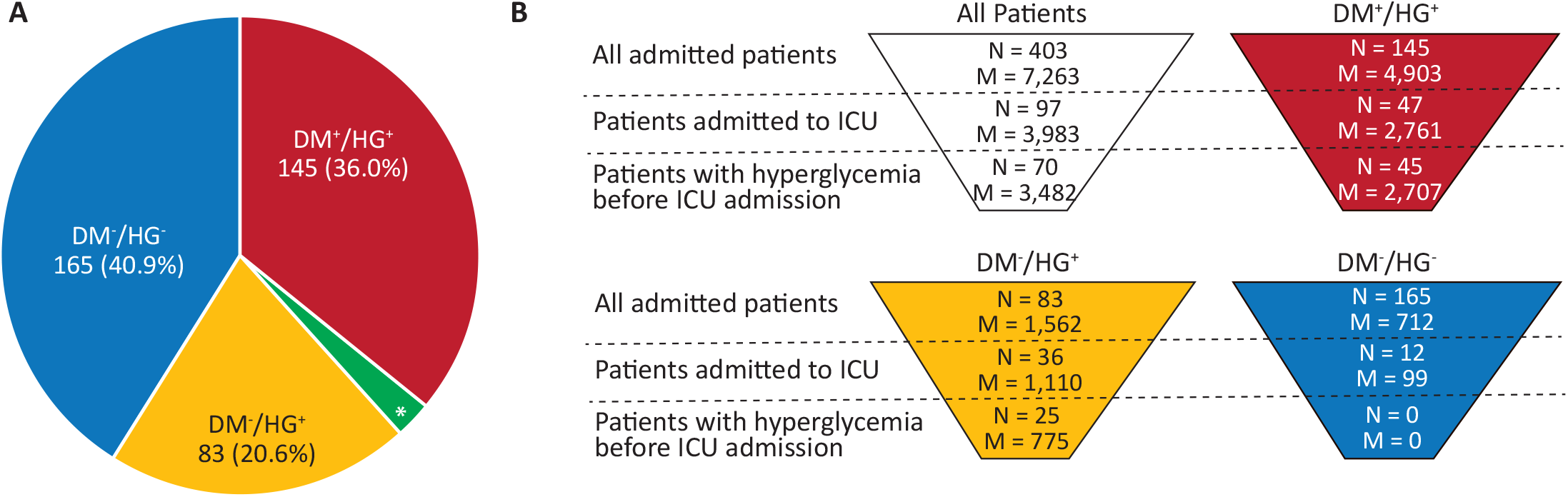
Distribution of study groups and blood glucose measurements in hospitalized COVID-19 patients. **(A)** The pie chart shows number (%) of patients in the color-coded study groups. These color-codes are consistently used throughout the rest of the paper. DM^+^/HG^+^, patients with diabetes and hyperglycemia; DM^+^/HG^-^, patients with diabetes but no hyperglycemia; DM^-^/HG^+^, patients with hyperglycemia who did not have diabetes; DM^-^/HG^-^, patients who had neither diabetes nor hyperglycemia **(B)** Funnel plots showing the distribution of patient subsets when considered in all study participants (funnel with white background); in the DM^-^/HG^+^ group (yellow background); in the DM^+^/HG^+^ group (red background); and in the DM^-^/HG^-^ group (blue background). N, number of patients; M, number of blood glucose measurements; *, DM^+^/HG^-^ group (N = 10 (2.5%)).

As shown in Figure 1B (White funnel), for the 403 patients included in this study, we had a total of 7,263 (average 18.0 per patient) BG measurements over the entire period of hospitalization. Of these, 3,983 BG measurements were on 97 patients who were admitted to ICU during hospital stay and 70/97 (72.2%) patients had their first hyperglycemia finding before ICU admission. Of the 83 patients in the DM^-^/HG^+^ group (Figure 1B, yellow funnel), 36 were admitted to ICU and 25/36 (69.4%) patients had their first hyperglycemia finding before ICU hospitalization. The average time (95% CI) from first hyperglycemia finding to ICU admission was 6.83 days (5.32 – 8.36 days) in all patients (n = 70) and 7.32 days (4.45 – 10.2 days) in the DM^-^/HG^+^ group (n = 25).

### Glycemia during hospitalization

Figure 2A shows the cubic spline smoothed, non-linear trends of glycemia in the study participants over the period of hospitalization according to the study groups. DM^-^/HG^-^ patients (blue curve) had well-maintained BG levels that stayed around 5.56 mmol/L throughout the first two-weeks of hospitalization with very little fluctuation. The DM^+^/HG^-^ patients (green curve) also mimicked the blue curve albeit with wider confidence bands and smaller number of data points. DM^+^/HG^+^ patients (red curve) demonstrated consistently high BG values with substantially larger fluctuations.

**Figure 2.**
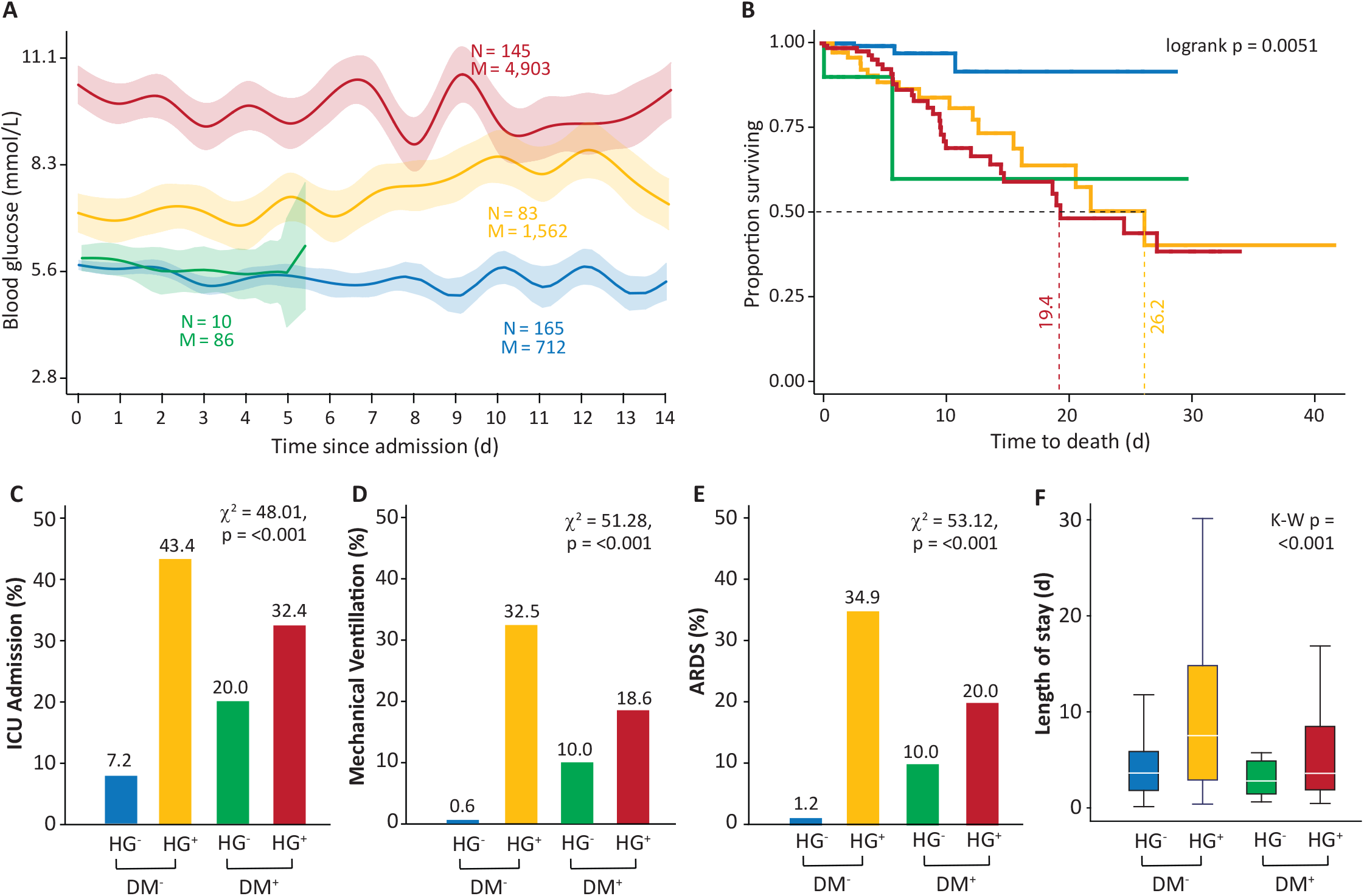
Glycemia trends and association of hyperglycemia with study outcomes in hospitalized COVID-19 patients. **(A)** Trends in glycemia over two-weeks following hospital admission for the diabetes- and hyperglycemia-based, color-coded study groups. N and M indicate number of patients and number of BG measurements, respectively. Shown in the plot for each study group are cubic spline-smoothed, non-linear glycemia trends obtained using generalized estimating equations. Thick lines show point estimates and light-colored areas show 95% confidence bands. **(B)** Kaplan-Meier plot for time to death in the color-coded study groups left-censored at the time of first detection of hyperglycemia. Median time to death is indicated using color-coded numbers and dashed vertical lines. Statistical significance for difference in survival curves was tested using the logrank test (indicated at the top-right corner). **(C-E)** Bar charts showing incidence of the indicated outcome (ICU admission, mechanical ventilation and ARDS in panels C, D, and E, respectively) across color-coded study groups. Association was tested using Pearson’s chi-square test which is shown on the top-right side of each panel. **(F)** Box plot showing the association of the color-coded study groups with length of stay in surviving patients. Median length of stay for each group is indicated by the white horizontal lines inside the colored boxes. Significance of association was tested using Kruskal-Wallis test (K-W P) the result of which is shown at the top-right side of panel F.

BG levels of the DM^-^/HG^+^ patients (yellow curve) showed an interesting pattern. For the first week, these patients had low average values that appeared to increase in the second week. Indeed, during the second week the DM^+^/HG^+^ and DM^-^/HG^+^ patients (red and yellow curve, respectively) showed overlapping confidence bands indicating no statistical difference in average BG levels. Consistent with these observations, the average coefficient of variation of BG values in the DM^+^/HG^+^(red curve), DM^+^/HG^-^(green curve), DM^-^/HG^+^(yellow curve) and DM^-^/HG^-^(blue curve) patients was 28.3%, 9.8%, 24.9% and 7.3%, respectively.

### Association of hyperglycemia and diabetes with mortality

We investigated the association of hyperglycemia with risk of hospital mortality in three steps. First, we ran a stepwise logistic regression model with a forward addition strategy that included a total of 59 covariates (all covariates are listed in the footnote to Table 2). Higher neutrophil percentage, older age, insulin therapy (as outpatient), hematuria, high serum globulin at admission, low platelet count and nasal congestion significantly associated covariates retained in this final model. In the second step of this analysis, we added hyperglycemia to the full model (total number of covariates 60) and found that hyperglycemia was retained in the final model with an OR of 14. In this model two new covariates (fever with chills and marijuana use) got added to the final model at the expense of the symptom of nasal congestion. The AUROC for models in step1 and step 2 was 0.8625 (95% CI 0.8105 – 0.9145) and 0.8979 (95% CI 0.8584 – 0.9374), implying a statistically significant improvement in prediction (0.0354, p = 0.0162). Lastly, to understand the association of hyperglycemia with and without diabetes, we replaced the covariate hyperglycemia with indicators for diabetes and hyperglycemia groups and used the DM^-^/HG^-^ patients as the reference group. The results (Table 2, step 3) showed that occurrence of hyperglycemia in patients with or without diabetes was significantly associated with hospital mortality. All other covariates retained in the final model in this step 3 were the same as those retained in step 2. This analytic strategy demonstrated that hyperglycemia with or without diabetes was an independent predictor of hospital mortality.

**Table 2.** Association of hyperglycemia and diabetes with the risk of death using stepwise logistic regression. Results shown are from final model for each scenario.

| Covariates | OR | 95% CI |
| --- | --- | --- |
| ***Step 1: using 59 covariates\**** |  |  |
| Differential neutrophil count** | 1.10 | 1.06 – 1.15 |
| Age | 1.05 | 1.02 – 1.08 |
| On insulin | 2.65 | 1.17 – 5.97 |
| Hematuria | 4.27 | 1.78 – 10.3 |
| Initial Serum globulin | 2.99 | 1.50 – 5.95 |
| Initial Platelet count | 0.99 | 0.99 – 1.00 |
| Nasal congestion | 9.06 | 1.33 – 61.9 |
| ***Step 2: Step 1 covariates + hyperglycemia*** |  |  |
| Differential neutrophil count** | 1.09 | 1.04 – 1.14 |
| Hyperglycemia | 14.0 | 3.47 – 56.3 |
| Age | 1.05 | 1.02 – 1.09 |
| Hematuria | 3.28 | 1.78 – 10.3 |
| Initial Serum globulin | 2.99 | 1.50 – 5.96 |
| Fever with chills | 5.69 | 1.51 – 21.5 |
| Marijuana use | 20.9 | 2.51 – 174.8 |
| Initial Platelet count | 0.99 | 0.99 – 1.00 |
| ***Step 3: Step 1 covariates + combination of diabetes and hyperglycemia*** |  |  |
| Differential neutrophil count** | 1.09 | 1.04 – 1.14 |
| Glycemic status |  |  |
| No-diabetes/no-hyperglycemia | Ref |  |
| No-diabetes/hyperglycemia | 21.94 | 4.04 – 119.0 |
| Diabetes/no-hyperglycemia | 5.97 | 0.32 – 111.8 |
| Diabetes/hyperglycemia | 17.06 | 3.46 – 84.1 |
| Age | 1.06 | 1.02 – 1.09 |
| Hematuria | 3.39 | 1.39 – 8.34 |
| Initial Serum globulin | 2.88 | 1.46 – 5.70 |
| Fever with chills | 6.10 | 1.62 – 23.0 |
| Marijuana use | 17.78 | 1.97 – 160.5 |
| Initial Platelet count | 0.99 | 0.99 – 1.00 |

\*, Results are from the final model retained after stepwise forward addition strategy. We included total 59 covariates – 4 sociodemographic variables (age, gender, ethnicity and black race), 12 symptoms (cough, cough with sputum, nasal congestion, headache, fever, fever with chills, shortness of breath, nausea/vomiting, diarrhea, myalgia, altered mental status and fatigue), 14 comorbidities (hypertension, diabetes, coronary artery disease, congestive heart failure, chronic obstructive pulmonary disease, asthma, other lung disease, chronic kidney disease, end-stage renal disease, chronic liver disease, cancer, ever smoker, alcohol consumption and qSOFA score) , 11 laboratory investigations (total white cell count, neutrophil count, lymphocyte count, platelet count, hemoglobin concentration, serum sodium, serum bicarbonates, serum creatinine, serum globulin, proteinuria, hematuria) 15 medications (insulin, angiotensin converting enzyme inhibitors, angiotensin receptor blockers, mineralocorticoid receptor antagonists, beta-blockers, other anti-hypertensive, statins, non-steroidal anti-
inflammatory drugs, aspirin, vitamin D, hydroxychloroquine, azithromycin, rivaroxaban, chemotherapy and steroids) and 3 substance use variables (marijuana, cocaine and heroin). Variables in the final model are shown in the order of entry into the model.
\*\*, used as a continuous variable and expressed as percentage
OR, odds ratio; CI, confidence interval; Ref, reference category

We assessed the robustness of this finding in several ways. First, the estimated ORs did not significantly differ between males and females (data not shown). Second, sensitivity analyses on patients with HbA1c data reaffirmed these results with a comparably high OR (13.0, Supplementary Table 1) and bootstrap CIs above unity. Third, the first detection of hyperglycemia was not influenced by the number of BG measurements ordered till that time point (adjusted OR 0.94, 95% CI 0.87 – 1.03). Lastly, we conducted a 10-fold cross-validation of the final model (Table 2, step 2); the results of which (Supplementary Table 2) showed a consistent prediction across folds (average 10-fold accuracy 0.7581) implying that the model did not overfit the data. Together, these results indicated that our observation of the association between hyperglycemia and mortality risk in hospitalized COVID-19 patients was robust to potential misclassification and indication biases.

### Hyperglycemia as an early indicator of mortality

To examine the clinical use of hyperglycemia as a predictor of mortality, we investigated whether detection of hyperglycemia early after admission can still provide a prognostic value. Average time from hospital admission to the first detection of hyperglycemia was 0.11 days (95% CI 0 – 0.27 days) in diabetes patients and 2.15 days (95% CI 1.46 – 2.83 days) in non-diabetes patients. Of the 228 patients with hyperglycemia, 177 (77.6%) were detected by the end of 24 hours and an additional 17 (total of 194, 85.1%) by the end of 48 hours. For this, we repeated the analyses shown in Table 2, step 2 by replacing the variable hyperglycemia with hyperglycemia detected during the first 24 hours (HG_24_) or 48 hours (HG_48_). We found (Supplementary Table 3) that both HG_24_ and HG_48_ were significant and independent predictor of mortality (HG 24 - OR 2.15, 95% CI 1.00 – 4.59, HG 48 OR - 3.31, 95% CI 1.44 – 7.62). Sensitivity analyses (Supplementary Table 1) in patients with HbA1c data suggested that while HG_24_ was marginally non-significant, HG_48_ was an independent predictor of mortality. Together, these results indicated that hyperglycemia was an early predictor of mortality in the study cohort.

### Association of hyperglycemia and diabetes with time to death

The Kaplan-Meier plot (Figure 2B) (left-censored at the first detection of hyperglycemia), showed 90% survival in the DM^-^/HG^-^ group (blue curve) while the DM^+^/HG^+^ patient group (red curve) rapidly progressed to death with a median survival time of 19.4 days. Interestingly, the DM^-^/HG^+^ group (yellow curve) overlapped the survival curve for the DM^+^/HG^+^ group with a median survival time of 26.2 days. Survival curves for the DM^+^/HG^+^(red curve) and DM^-^/HG^+^(yellow curve) patients crossed each other towards the end of follow-up indicating comparable survival. These results showed that the hyperglycemia and diabetes-based patient groups were significantly associated (logrank p=0.005) with survival in hospitalized COVID-19 patients.

### Association of hyperglycemia and diabetes with secondary outcomes

Consistent with the results presented thus far, we observed that the patients in the DM^+^/HG^+^(red bars) and DM^-^/HG^+^(yellow bars) groups had significantly high rates of ICU admission, mechanical ventilation and development of ARDS (Figure 2, panels C-E). These results were replicated when restricted to patients with HbA1c data (Supplementary Table 4). HG_24_ and HG_48_ were significantly associated with all the three outcomes (Supplementary Table 5) affirming the prognostic use of early hyperglycemia. Lastly, we replicated these analyses in the subset of patients where the first detection of hyperglycemia preceded the specific outcome. The results of these analyses (Supplementary Table 6) corroborated with those shown in Figure 2 (panels C-E). Further, in the surviving patients, those belonging to the DM^-^/HG^+^ group (yellow box in Figure 2F) had significantly long median length of hospital stay (7.6 days) as compared to the other three groups (ranging from 2.9 to 3.8 days) (Kruskal-Wallis p <0.001).

## DISCUSSION

The prevalence of hyperglycemia in our study of hospitalized COVID-19 patients was 57% which is higher than the 38-40% prevalence reported in non-COVID-19 patients.[18] Our analyses have uncovered hospitalized non diabetic COVID-19 patients with hyperglycemia as a subgroup (21%) that is associated with a high risk of death; have a rapid progression to death; and have a generally challenging clinical course. Diabetes patients are a known high-risk group in COVID-19 disease.[2] While our results support this view, they also imply that it may be more informative to focus on glycemic status as an indicator of the clinical course of COVID-19 patients. Focusing on hyperglycemia, as shown by results in Table 2 and Figure 2, can potentially inform a clinician early and accurately about the anticipated disease course. Our results also point toward the possibility of using hyperglycemia within the first 48 hours of admission as an independent predictor of COVID-19 prognosis. It is noteworthy that our definition of hyperglycemia is biased towards picking up hyperglycemia early during disease.

We and others have previously demonstrated that the degree of hyperglycemia in critically ill patients without diabetes plays a significant prognostic role in predicting hospital mortality.[8, 19, 20] Indeed, Max Harry Weil, father of critical care medicine, knew by 1973 that in critically ill patients, “Elevation of blood sugar reflects secretion of increased amounts of catecholamines from the adrenal medulla.”[21] Epinephrine-induced phosphorylation of the insulin receptor reduces its tyrosine kinase activity[22] and causes prompt and prolonged inhibition of pancreatic insulin secretion.[23] Thus, early appearance of hyperglycemia in non-diabetic COVID-19 patients likely signals increased systemic stress.

However, this prognostic role of hyperglycemia varies by the primary cause of critical care such that primary diagnoses like trauma, coma and neurological diseases are especially prone to high likelihood of adverse outcomes associated with hyperglycemia.[8] Our results need to be viewed in the light of the emerging literature on association of hyperglycemia with COVID-19 prognosis. Studies have shown that pre-existing diabetes [2], newly detected diabetes [24], prediabetes [25], uncontrolled hyperglycemia (≥2 BG values of ≥10 mMol/L[3]) or fasting BG ≥7 mmol/L [26] are significant determinants of COVID-19 prognosis. Our study adds to these findings the observation that hyperglycemia detected early after hospitalization in patients without a history of diabetes can also independently predict the disease course in COVID-19 patients.

In this regard, hyperglycemia could be causally involved in the development of cytokine storm[14, 27, 28] and severe lung pathology in critically ill COVID-19 patients. This reasoning arises from recent seminal work of Hoepel *et al*. [29], which begins with observations that (1) onset of severe pulmonary disease coincides with adaptive immunity production of IgG ∼10 days after initial symptom onset and (2) there is increased glycosylation of the Fc of anti-Spike IgG compared to total IgG in sick COVID-19 patients. These investigators showed that immune complexes of COVID-19 Spike protein with anti-Spike IgG from critically ill COVID-19 patients elicited dose-dependent production of inflammatory cytokines by human M2 macrophages. In a further *in vitro* model with human pulmonary artery endothelial cells and platelets, these Spike-IgG immune complexes and macrophages induced long-lasting endothelial disruption, increased platelet adhesion to the endothelial cells, and release of von Willebrand Factor. The same experiment using immune complexes of recombinant IgG-Spike-protein elicited much less pro-inflammatory macrophage response; however, after glycosylation of the recombinant anti-Spike IgG, Spike-IgG immune complexes again drove the macrophage inflammatory responses. Specific blockade of the macrophage Fcγ2 receptor strongly inhibited the inflammatory response to Spike-IgG immune complexes. We note that this report of platelet activation induced by glycosylated-immune complex stimulation of macrophages is consistent with platelet hyperactivity in COVID-19 patients[30] and autopsy histopathology identifying platelet-fibrin microthrombi in the lungs[31]. It has become clear that a key characteristic that determines IgG pathogenicity is glycosylation on the IgG Fc tail.[32] Elevated HbA1c from hyperglycemia is associated with pro-inflammatory glycosylation of IgG Fc in both Type 1 and Type 2 diabetics.[33, 34] Even in non-diabetics HbA1c increases with age[35], and elevated HbA1c predicts a more difficult course of COVID-19.[36]

The above observations suggest a causal hypothesis linking hyperglycemia, excessive macrophage activation, respiratory distress, and hypercoagulopathy in the pathogenesis of severe COVID-19. COVID-19 suppresses both innate immunity by obstructing interferon production[37-39] and also adaptive immunity by infecting lymphocytes[40], inducing their apoptosis and lymphopenia, and markedly limiting T-cell response.[41] This leaves an immune response involving predominantly MHC1 antigenic epitopes, Natural-Killer cells, and macrophage activation.[42] Plasma cells do eventually respond with IgG; however, with hyperglycemia, anti-Spike Fc-glycosylated-IgG produces immune complexes that stimulate the pathology of full-blown macrophage activation syndrome (MAS).[43] This results in excessive platelet activation and hypercoagulability with compromised microperfusion together with pulmonary endothelial fluid leakage and severe respiratory distress syndrome with progressive hypoxia leading to death.

We have observed the appearance of hyperglycemia in non-diabetic COVID-19 patients who are much more likely to progress to severe disease. We suggest this is an early marker of a stress response that results in amplification of the pathophysiology outlined above. Close and perhaps continuous monitoring of blood glucose in hospitalized COVID-19 patients could provide clinicians with early recognition of this risk. Future studies are needed to confirm or refute these mechanistic hypotheses.

Our study has some limitations. First, this is an observational study of hospitalized COVID-19 patients and is thus prone to all the limitations of observational studies. For instance, causative association cannot be established from our results. While our analyses attempted to ensure that hyperglycemia preceded the outcomes of interest, the possibility of a temporal bias cannot be refuted. Second, our study does not include any practice changes based on hyperglycemia detection. However, our results suggest that a closer clinical scrutiny of COVID-19 patients based on glycemic status may provide additional insights into their clinical course. Studies in future need to specifically address these hypotheses. Third, although we have provided a coherent model of the pathophysiology, whether hyperglycemia in COVID-19 patients is consequential or coincidental to disease biology is currently unknown and cannot be surmised from our results. Studies are needed to specifically understand the biology of hyperglycemia in COVID-19 patients. Fourth, since the BG data were retrospectively derived from the source of blood sample, its relation to fasting status remains unknown in this study. This heterogeneity of BG sampling could have biased our OR estimates. Despite this potential measured and unmeasured confounding, our results from sensitivity analyses indicate that our interpretations are likely to have been minimally influenced by confounding due to BG sampling. Fifth, the likelihood of undiagnosed diabetes at admission was 4% and 10% in the no-diabetes/no-hyperglycemia and no-diabetes/hyperglycemia groups, respectively. We evaluated, through sensitivity analyses, the influence of this misclassification on our interpretations. While the sensitivity analyses demonstrated the robustness of our inferences, larger multicentric studies are needed before the findings can be generalized.

Nevertheless, we present evidence in favor of hyperglycemia as an early marker of clinical deterioration in COVID-19 patients. Hyperglycemia as defined in this study is mostly inclusive of and incremental to known diabetes status both in terms of prevalence and its association with mortality. A shift of focus from diabetes to hyperglycemia can enable early identification of patients at risk for poor outcomes and thus improve risk stratification of COVID-19 patients.

## Data Availability

The data used in this study represent confidential patient-level information that cannot be shared publicly.

## ACKNOWLEDGMENTS

None of the authors have a conflict of interest to disclose. H.K., A.M.A. and M.M. conceptualized the study. H.K. and M.M. conducted statistical analyses and wrote the first draft of the manuscript. A.M.A., M.A., J.V., A.R.A., J.P.R., A.J.J., M.I., S.Z., R.P.M. and P.H. contributed to data collection and critical revision of the manuscript. B.C.W. provided the pathophysiological foil to the inference and wrote the related parts. All authors read and approved the final version of the manuscript. Funding – None.

