## Supplementary Material for "ASSOCIATION OF HYPERGLYCEMIA WITH HOSPITAL MORTALITY IN NONDIABETIC COVID-19 PATIENTS: A COHORT STUDY"

**Supplementary Table 1. Sensitivity analyses for the association of hyperglycemia with mortality in patients with available HbA1c data***

| **Covariates** | **OR** | **Bootstrap**  **95% CI** |
| --- | --- | --- |
| ***Hyperglycemia*** | | |
| Differential neutrophil count** | 1.09 | 1.04 – 1.15 |
| Hyperglycemia | 13.01 | 3.49 – 48.4 |
| Age | 1.04 | 1.00 – 1.08 |
| Hematuria | 4.37 | 1.42 – 13.5 |
| Initial Serum globulin | 2.87 | 1.11 – 7.44 |
| Initial Platelet count | 0.99 | 0.99 – 1.00 |
| ***Hyperglycemia within 24 hours of admission*** | | |
| Differential neutrophil count** | 1.10 | 1.05 – 1.16 |
| HG_24_ | 2.91 | 0.86 – 9.87 |
| Age | 1.03 | 0.99 – 1.07 |
| Hematuria | 4.77 | 1.54 – 14.8 |
| Initial Serum globulin | 2.76 | 1.09 – 6.94 |
| Initial Platelet count | 0.99 | 0.99 – 1.00 |
| ***Hyperglycemia within 48 hours of admission*** | | |
| Differential neutrophil count** | 1.10 | 1.04 – 1.15 |
| HG_48_ | 4.95 | 1.07 – 23.0 |
| Age | 1.03 | 0.99 – 1.07 |
| Hematuria | 4.61 | 1.57 – 13.5 |
| Initial Serum globulin | 2.95 | 1.10 – 7.84 |
| Initial Platelet count | 0.99 | 0.99 – 1.00 |

*Compared to the results shown in Table 2, step 2, the following two variables were not retained in the final model here – fever with chills and marijuana use

**, used as a continuous variable and expressed as percentage

**Supplementary Table 2. Ten-fold cross validation of the final model (from Step 2, Table 2)**

| Training set | Validation set | Training AUROC | Validation AUROC |
| --- | --- | --- | --- |
| Folds 2-10 | Fold 1 | 0.8037 | 0.7854 |
| Folds 1, 3-10 | Fold 2 | 0.8031 | 0.8243 |
| Folds 1-2, 4-10 | Fold 3 | 0.8113 | 0.7292 |
| Folds 1-3, 5-10 | Fold 4 | 0.8313 | 0.5778 |
| Folds 1-4, 6-10 | Fold 5 | 0.7809 | 0.7396 |
| Folds 1-5, 7-10 | Fold 6 | 0.8063 | 0.9487 |
| Folds 1-6, 8-10 | Fold 7 | 0.7932 | 0.7692 |
| Folds 1-7, 9-10 | Fold 8 | 0.8036 | 0.8235 |
| Folds 1-8, 10 | Fold 9 | 0.8181 | 0.7667 |
| Folds 1-9 | Fold 10 | 0.8187 | 0.6171 |
| Average accuracy | | 0.8070 | 0.7581 |

AUROC, Area Under the Receiver Operating Characteristic Curve

**Supplementary Table 3. Association of time-dependent detection of hyperglycemia with mortality**

| **Covariates** | **OR** | **95% CI** |
| --- | --- | --- |
| ***Hyperglycemia within 24 hours of admission*** | | |
| Differential neutrophil count* | 1.10 | 1.05 – 1.15 |
| HG_24_ | 2.15 | 1.00 – 4.59 |
| Age | 1.05 | 1.02 – 1.08 |
| Hematuria | 4.41 | 1.90 – 10.2 |
| Initial Serum globulin | 2.94 | 1.52 – 5.66 |
| Fever with chills | 3.67 | 1.18 – 11.5 |
| Marijuana use | 7.36 | 1.15 – 47.0 |
| Initial Platelet count | 0.99 | 0.99 – 1.00 |
| ***Hyperglycemia within 48 hours of admission*** | | |
| Differential neutrophil count* | 1.09 | 1.05 – 1.14 |
| HG_48_ | 3.31 | 1.44 – 7.62 |
| Age | 1.05 | 1.02 – 1.08 |
| Hematuria | 4.22 | 1.81 – 9.84 |
| Initial Serum globulin | 3.02 | 1.55 – 5.88 |
| Fever with chills | 3.87 | 1.18 – 12.6 |
| Marijuana use | 8.81 | 1.33 – 58.3 |
| Initial Platelet count | 0.99 | 0.99 – 1.00 |

*, used as a continuous variable and expressed as percentage

**Supplementary Table 4. Association of hyperglycemia with secondary outcomes in patients in whom HbA1c values were available and who were reclassified as diabetes if HbA1c was ≥6.5%**

| Outcome | DM^+^/HG^+^  (n = 151)  N (%) | DM^+^/HG^-^  (n = 11)  N (%) | DM^-^/HG^+^  (n = 41)  N (%) | DM^-^/HG^-^  (n = 76)  N (%) | p |
| --- | --- | --- | --- | --- | --- |
| ICU admission | 49 (32.5) | 2 (18.2) | 17 (41.5) | 4 (5.3) | <0.001 |
| Mechanical ventilation | 30 (19.9) | 1 (9.1) | 12 (29.3) | 0 (0.0) | <0.001 |
| ARDS | 32 (21.2) | 1 (9.1) | 13 (31.7) | 1 (1.3) | <0.001 |

DM^+^/HG^+^, patients with diabetes and hyperglycemia; DM^+^/HG^-^, patients with diabetes but no hyperglycemia; DM^-^/HG^+^, patients with hyperglycemia who did not have diabetes; DM^-^/HG^-^, patients who had neither diabetes nor hyperglycemia

**Supplementary Table 5. Association of time-dependent detection of hyperglycemia with secondary outcomes in patients without diabetes.**

| Outcome | Incidence (%) in  DM^-^/HG^+^ | Incidence (%) in  DM^-^/HG^-^ | χ^2^ | p |
| --- | --- | --- | --- | --- |
| ***Hyperglycemia within 24 hours of admission*** | | | | |
| ICU admission | 40.0 | 14.8 | 15.01 | <0.001 |
| Mechanical ventilation | 28.9 | 7.4 | 17.00 | <0.001 |
| ARDS | 33.3 | 7.9 | 21.82 | <0.001 |
| ***Hyperglycemia within 48 hours of admission*** | | | | |
| ICU admission | 38.9 | 13.9 | 16.88 | <0.001 |
| Mechanical ventilation | 27.8 | 6.7 | 18.74 | <0.001 |
| ARDS | 31.5 | 7.2 | 22.74 | <0.001 |

DM^-^/HG^+^, patients with hyperglycemia who did not have diabetes; DM^-^/HG^-^, patients who had neither diabetes nor hyperglycemia; χ^2^, chi square value; p, significance value

**Supplementary Table 6. Sensitivity analyses of the association of diabetes/hyperglycemia groups with secondary outcome by restricting to occurrence of hyperglycemia before the outcome**

| Outcome | DM^+^/HG^+^ | DM^+^/HG^-^ | DM^-^/HG^+^ | DM^-^/HG^-^ | χ^2^(df), p |
| --- | --- | --- | --- | --- | --- |
| ICU Admission | 45 (31.5%) | 0 (0.0%) | 25 (34.7%) | 0 (0.0%) | 0.23 (1), 0.631 |
| Mechanical ventilation | 26 (18.1%) | 0 (0.0%) | 16 (22.2%) | 0 (0.0%) | 0.53 (1), 0.466 |
| ARDS | 29 (20.0%) | 1 (10.0%) | 29 (34.9%) | 2 (1.2%) | 53.12 (3), <0.001 |

DM^+^/HG^+^, patients with diabetes and hyperglycemia; DM^+^/HG^-^, patients with diabetes but no hyperglycemia; DM^-^/HG^+^, patients with hyperglycemia who did not have diabetes; DM^-^/HG^-^, patients who had neither diabetes nor hyperglycemia; χ^2^, chi square value; df, degree of freedom; p, significance value
